# Enhancing Thyroid Nodule Assessment: Leveraging Contrast-Enhanced Ultrasonography as a Screening Modality - A Meta-Analysis

**DOI:** 10.1101/2024.08.11.24304174

**Authors:** Dev Desai, Maria Eleni Malafi, Hetvi Shah, Aneri Parikh, Abhijay B. Shah, Vismit Gami, Parth Gupta

## Abstract

**Introduction:** CEUS has become an emerging radio diagnostic technique of modern times. The use of these contrasts offers a way better alternative than materials that cause radiation. Thyroid nodules are notorious for their effect on normal physiology and the routine best diagnostic modality apart from biopsy is Radioactive Iodine.

**Aim:** To conduct a diagnostic test accuracy meta-analysis to understand the role of CEUS in diagnosing thyroid nodules.

**Methodology:** According to Prisma guidelines, literature on the topic was found using the Keywords CEUS, Thyroid Nodule, and Radioactive Iodine. Two independent reviewers conducted a quality check on the papers and decided on the studies that should be included. Any discrepancies were solved by a third reviewer. Meta Disc, Review Manager, and Excel were used to analyze the extracted data from the selected studies as per the inclusion and exclusion criteria. Biopsy was taken as a Reference Gold Standard.

**Result:** A total of 47 RCTs with 5,527 patients were selected for the study. The pooled sensitivity of CEUS is 0.87, with a CI of 95% in a range of 0.86 to 0.88. The specificity of CEUS is 0.84, with a CI of 95% in a range of 0.82 to 0.85. The summary of the ROC curve shows that the area under the curve for CEUS was 0.9292 and the overall diagnostic odds ratio (DOR) was 40.59.

**Conclusion:** It can be concluded from the results that CEUS can be used as a Screening tool for high suspicion groups but it is still not a perfect test. The newer generation of Contrasts may yield higher accuracy but for the currently available contrasts, Biopsy remains the best tool for a definitive and accurate diagnosis.

## INTRODUCTION

A thyroid nodule is an atypical mass or cluster of cells on the thyroid gland. They are frequently encountered, typically noncancerous (benign), and often asymptomatic. In rare instances, they may be cancerous. Over 90% of nodules identified in adults are not cancerous (benign), yet they could indicate thyroid cancer in about 4.0% to 6.5% of instances. (1). Certain nodules can disrupt the thyroid glands’ hormone production, leading to signs of either hypothyroidism (a thyroid gland that’s not producing enough hormones) or hyperthyroidism (an overly active gland). Numerous nodules remain asymptomatic until they reach a size that impacts nearby tissues or becomes visible on the neck’s surface. Depending on the nodule’s type and origin, symptoms can encompass: difficulty swallowing, hoarseness, neck discomfort, rapid weight loss, cold intolerance, irregular pulse, fatigue, dry skin, facial edema, etc(2). The initial evaluation of a patient with a suspected thyroid nodule should be comprehensive, involving all aspects of a holistic investigative approach. Thyroid USG serves multiple purposes that indicate the benefits of wider application of this modality towards better diagnosis and management of the underlying condition. This ultrasound serves multiple purposes, including confirming the presence of the nodule, examining for any additional nodules, evaluating cervical lymph nodes, and assessing for any suspicious ultrasound(3). Ultrasound holds significant promise for assisting in distinguishing between malignant and benign thyroid nodules, but it is plagued by interpretational challenges, and its precision remains limited(4). Contrast-enhanced ultrasound involves the use of an intravenous substance containing microbubbles. This contrast agent assists healthcare providers in visualizing blood flow within your organs and blood vessels, aiding in the detection of a wide range of diseases and medical conditions(5). The purpose of this study is to ascertain the impact, merits, and demerits of CEUS as a diagnostic modality for thyroid nodules.

## METHODOLOGY

### DATA COLLECTION

For the collection of the data, a search was done by two individuals using PubMed, Google Scholar, and Cochrane Library databases for all relevant literature. Full - Text Articles written only in English were considered.

The medical subject headings (MeSH) and keywords, ‘Thyroid Nodule’, ‘Contrast-Enhanced Ultrasound’, ‘Benign’, ‘Malignant’, and ‘Prognosis’ were used. Original articles, reviews, and meta-analyses were scanned for additional articles.

### INCLUSION AND EXCLUSION CRITERIA

Titles and abstracts were screened and duplicates and references were removed. References to relevant articles were checked for additional articles. Literature patient information and test results were selected.

We searched for articles that showed more accurate indications that the method assessed was CEUS.

The inclusion criteria were as follows: (1) studies providing information on accurate diagnosis by CEUS; (2) research published in English; (3) Studies comparing CEUS with the gold standard (biopsy) as a diagnostic method for cases of thyroid nodules.

Exclusion criteria were: (1) articles not in full text, (2) unpublished articles, and (3) articles in other languages.

### DATA EXTRACTION

Each article was reviewed independently by two reviewers. Each article was analyzed for the number of patients, their age, their occupation and the incidence of pre-specified complications. Further discussion or discussion with the author and a third party was used to resolve the issue. Study quality was assessed using a modified Jadad score. Finally, according to PRISMA, 47 RCTs on the use of CEUS with a total of 5527 patients were considered for screening..

### ASSESSMENT OF STUDY QUALITY

Using the QualSyst tool, two authors independently assessed the efficacy of each included study. There are 10 questions in this test, each question has a score between 0 and 2, and 20 questions is the maximum mark possible. Two authors independently assessed each article based on the above criteria. Interobserver agreement on study selection was determined using Cohen’s Kappa (K). We also used the Cochrane tool to assess risk of bias for RCTs. We are not responsible for missing or unclear data. There is no funding for data collection and analysis..

### STATISTICAL ANALYSIS

RevMan (Review Manager, version 5.3), SPSS (Statistics for Social Sciences, version 20), Google Sheets and Excel in Stata 14 were used to perform the statistical analysis. The data were obtained and entered into the analysis software [21]. Fixed or random effects models were used to estimate sensitivity, specificity, positive predictive value (PPV), odds ratio (DOR) and relative risk (RR) with 95% confidence intervals. to examine clinically important outcomes (CI). Diagnostic accuracy and the Yonden index were calculated for each product. The sensitivity and specificity of individual probes were plotted on forest plots and receiver operating characteristic (ROC) plots. A forest plot and Fagan’s nomogram were used to show the sensitivity and specificity of different transcripts.

### BIAS STUDY

The risk of bias was evaluated by using QUADAS-2 analysis. This tool includes 4 domains Patient selection, Index test, Reference standard, Flow of the patients, and Timing of the Index tests.

## RESULT

**Figure 2:**
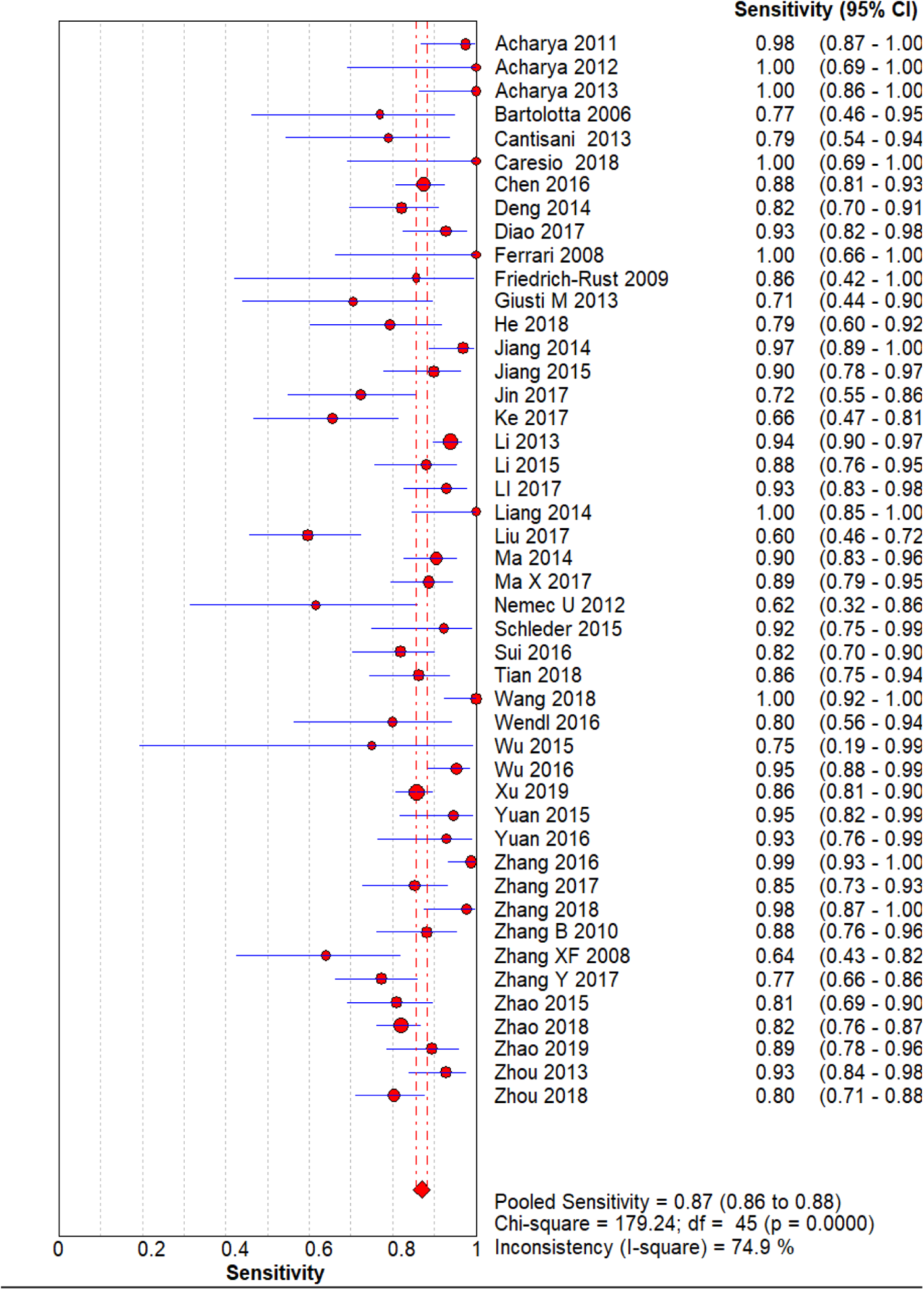
The forest chart summary for pooled sensitivity values of thyroid nodules.

**Figure 3:**
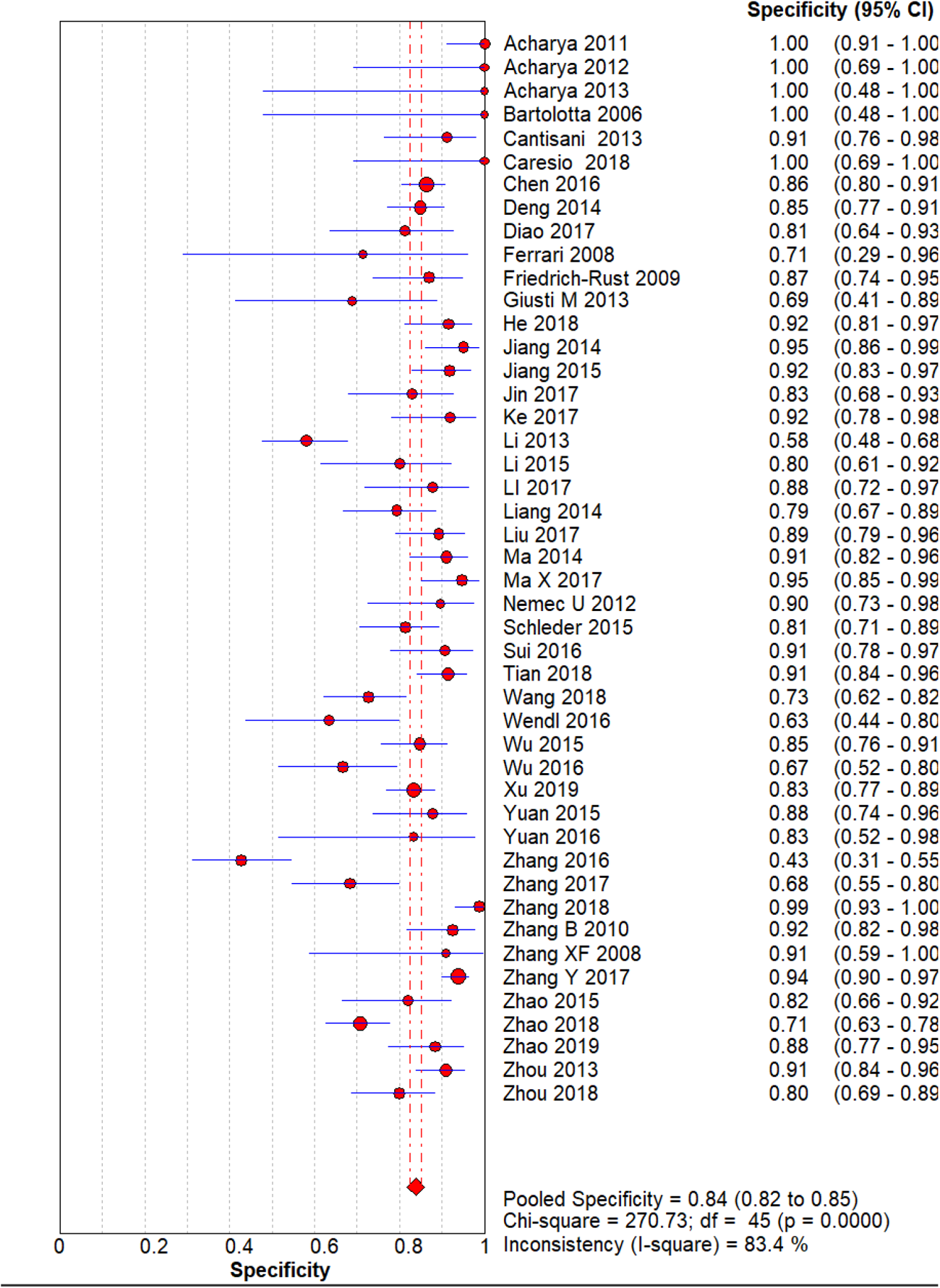
The forest chart summary for pooled specificity values of thyroid nodules.

**Figure 4:**
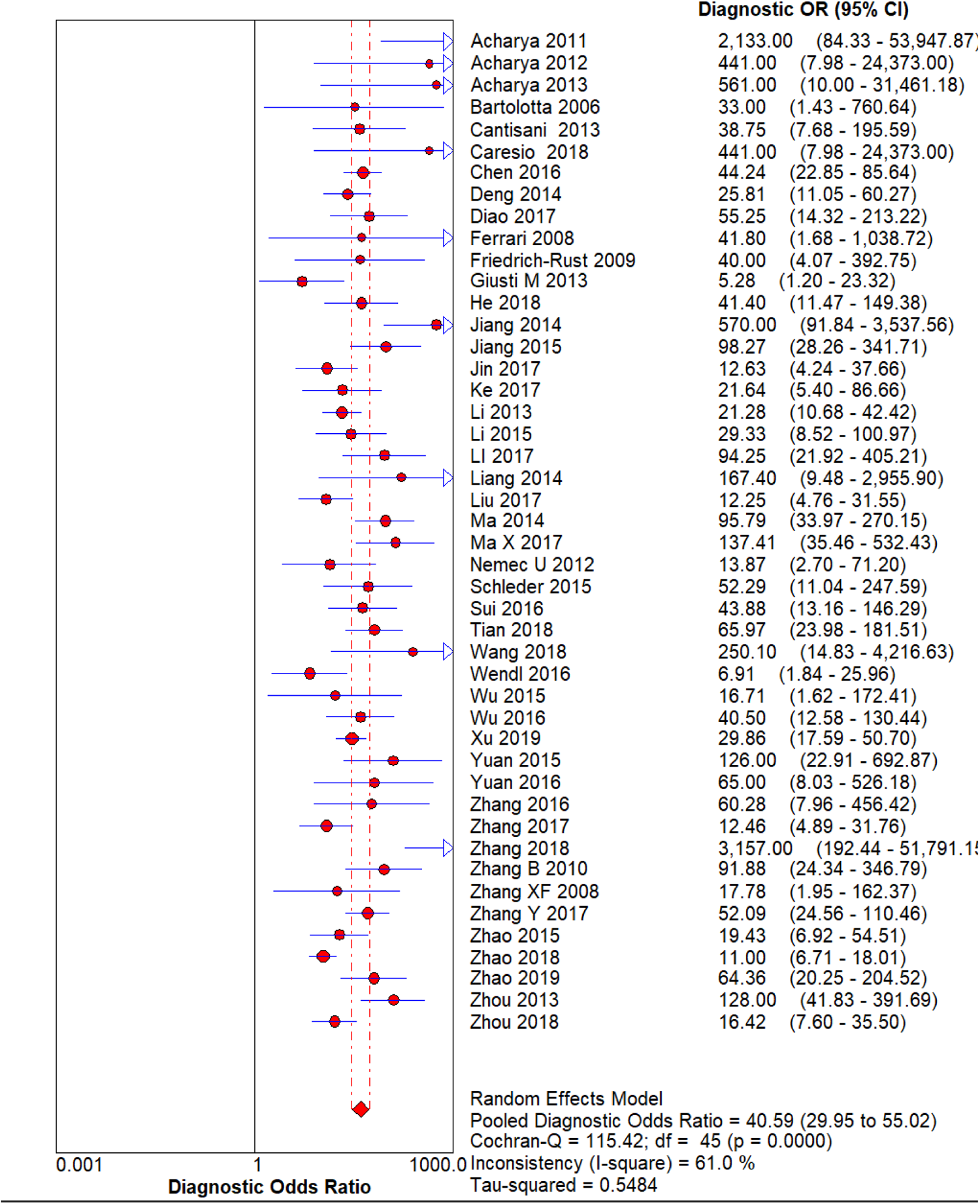
The forest chart summary for pooled diagnostic odds ratio of thyroid nodules.

**Figure 5:**
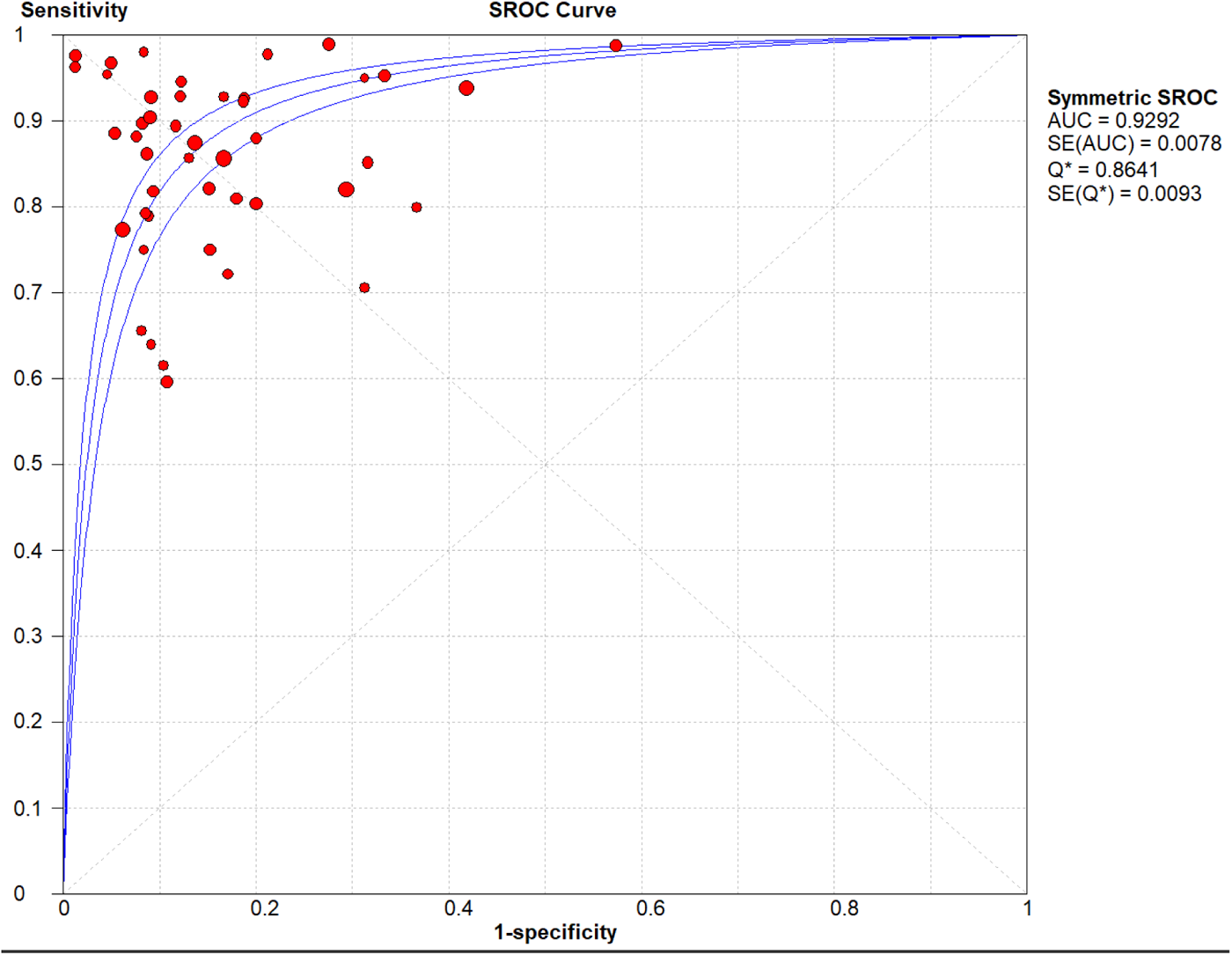
The SROC plot summary for CEUS for thyroid nodules.

**Figure 6:** Funnel Plot for CEUS for thyroid nodules.

### CEUS vs HISTOPATHOLOGY for thyroid nodules

Here, Table 1 describes all the descriptions of papers used for the CEUS vs Histopathology study. All the results described above, in the forest chart (figures 2 and 3), the comparison of the sensitivity and specificity of different papers can be observed. The same is illustrated in the SROC curve. (Figure 5). A total of 47 RCTs with 5,527 patients were selected for the study, out of which 12 studies showed sensitivity at or above 95%, and 8 studies showed specificity at or above 95%. 6 studies showed both sensitivity and specificity, to be at or over 95%. The value of True Positive (TP) was 2,306, that of True Negative (TN) was 2,414, that of False Positive (FP) was 464, and that of False Negative (FN) was 343. The pooled sensitivity of CEUS is 0.87, with a CI of 95% in a range of 0.86 to 0.88. The specificity of CEUS is 0.84, with a CI of 95% in a range of 0.82 to 0.85.

The summary of the ROC curve is described in Figure 5. It shows that the area under the curve for CEUS was 0.9292 and the overall diagnostic odds ratio (DOR) was 40.59.

**Figure 7:**
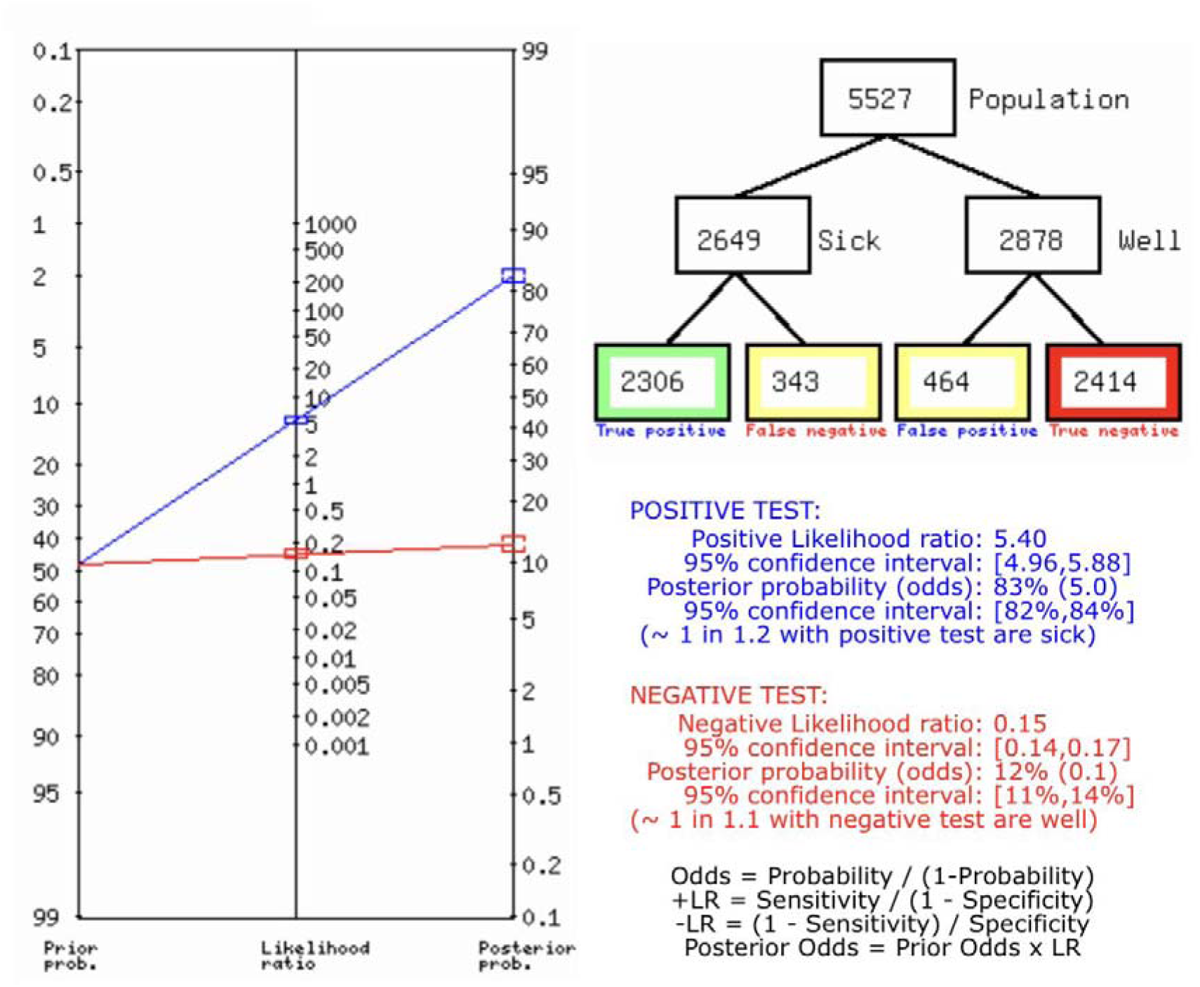
Fagan’s Analysis for CEUS vs HISTOPATHOLOGY for thyroid nodules.

Figure 7 describes the summary of Fagan plot analysis for all the studies considered for CEUS vs Histopathology for diagnosis of thyroid nodules, showing a prior probability of 48% (0.9); a Positive Likelihood Ratio of 5.40; a probability of post-test 83% (5.0); a Negative Likelihood Ratio of 0.15, and a probability of post-test 12% (0.1).

**Figure 8:**
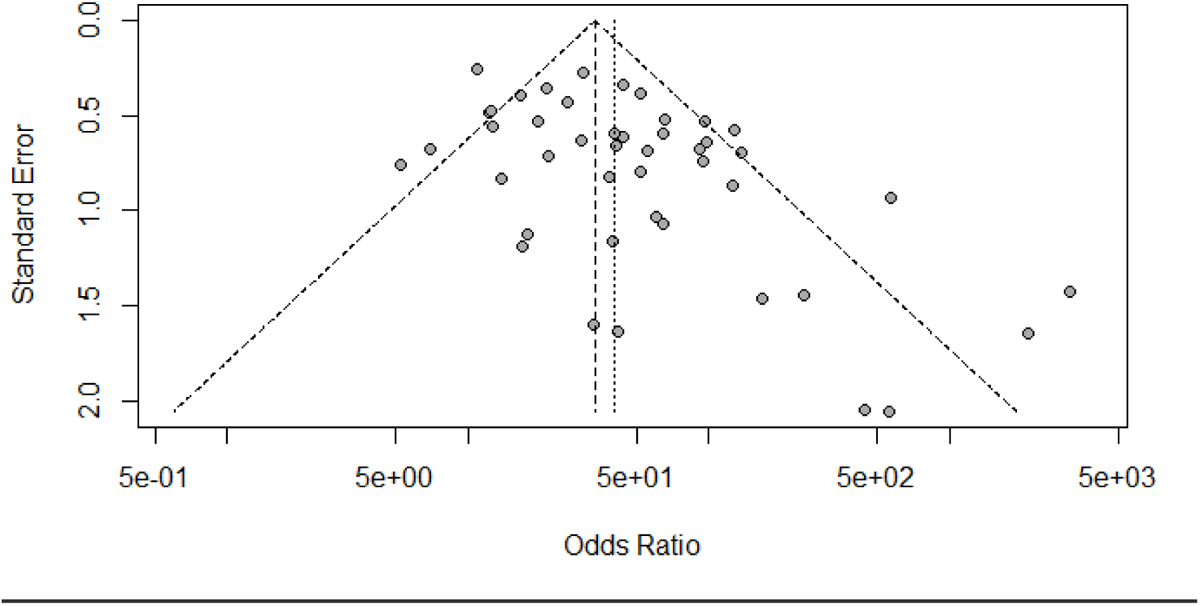
Deek’s Funnel Plot for CEUS vs HISTOPATHOLOGY for thyroid nodules.

## BIAS STUDY

|  | Patient Selection | Index Test | Reference Standard | Flow and Timing |  | Patient Selection | Index Test | Reference Standard |
| --- | --- | --- | --- | --- | --- | --- | --- | --- |
| Bozbora 2002 | low | low | low | low |  | high | low | low |
| Gannon 2018 | low | low | low | high |  | low | low | low |
| Jul 2009 | low | unclear | low | low |  | low | low | low |
| Li 2016 | high | low | low | low |  | low | low | low |
| Lin 2009 | low | low | unclear | low |  | low | low | low |
| Ma 2014 | unclear | high | low | low |  | low | low | low |
| Palaniappan 2016 | low | low | high | low |  | high | low | low |
| Stacul 2007 | low | low | low | low |  | high | low | low |
| Acharya 2011 | low | low | high | unclear |  | low | unclear | low |
| Acharya 2012 | low | unclear | low | low |  | low | low | low |
| Bartolotta 2006 | unclear | low | unclear | low |  | high | low | low |
| Caresio 2018 | low | low | high | low |  | low | low | low |
| Chen 2016 | low | low | unclear | low |  | low | low | low |
| Ferrari 2008 | low | unclear | low | unclear |  | unclear | low | low |
| Friedrich-Rust 2009 | low | low | high | low |  | low | low | low |
| Giusti M 2013 | low | low | low | high |  | low | low | low |
| He 2018 | low | high | low | low |  | low | high | low |
| Jiang 2014 | low | low | high | low |  | low | low | low |
| Jiang 2015 | unclear | low | low | low |  | high | low | low |
| Ke 2017 | low | unclear | low | low |  | low | low | low |
| Liu 2017 | low | low | low | low |  | high | low | low |
| Ma 2014 | low | low | low | high |  | low | high | low |
| Nemec U 2012 | low | low | low | high |  | low | low | low |
| Sui 2016 | low | high | low | low |  | low | low | low |
| Tian 2018 | low | low | unclear | low |  | low | low | low |
| Wang 2018 | high | low | low | low |  | low | low | low |
| Wu 2015 | low | unclear | low | low |  | low | low | low |
| Xu 2019 | low | low | unclear | low |  | high | low | low |
| Yuan 2016 | low | low | high | low |  | low | low | low |
| Zhang 2016 | low | low | high | low |  | low | low | low |
| Zhang 2018 | low | unclear | low | low |  | low | low | low |
| Zhang B 2010 | unclear | low | low | low |  | high | low | low |
| Zhang XF 2008 | low | low | low | low |  | low | unclear | low |
| Zhang Y 2017 | high | unclear | low | low |  | low | low | low |
| Zhao 2018 | low | low | high | low |  | low | low | low |
| Zhao 2019 | low | low | unclear | low |  | low | low | low |
| Zhou<br>2013 | low | high | low | low |  | unclear | low | low |
| Zhou<br>2018 | low | low | low | unclear |  | high | low | low |
| Basharat<br>2011 | low | low | high | low |  | low | low | low |
| Deandreis<br>2012 | low | low | low | high |  | low | low | low |
| Lin 2009 | low | high | low | low |  | low | high | low |
| Lumachi<br>2004 | low | high | low | low |  | unclear | low | low |
| Muñoz<br>Pérez<br>2013 | low | low | low | low |  | low | low | low |
| Piccardo<br>2016 | low | low | unclear | low |  | high | low | low |
| Riazi<br>2014 | low | unclear | low | low |  | unclear | low | low |
| Chen<br>2017 | low | low | low | low |  | low | low | low |
| Diao<br>2016 | unclear | low | high | low |  | low | low | low |
| He 2020 | low | low | low | unclear |  | low | low | low |
| Li 2017 | low | high | low | low |  | low | high | low |
| Li 2020 | low | low | low | high |  | unclear | low | low |
| Yang<br>2017 | low | low | high | low |  | low | low | low |
| Zhao<br>2019 | low | low | low | high |  | low | unclear | low |
| Zhu 2018 | low | low | low | high |  | low | low | low |
| total papers | 67 |  |  |  |  |  |  |  |
| Risk of Bias | Low | Unclear | High |  |  | Low | Unclear | High |
| Patient Selection | 56 | 6 | 5 |  | Patient Selection | 51 | 6 | 10 |
| Index test | 47 | 8 | 12 |  | Index test | 54 | 8 | 7 |
| Reference Standard | 48 | 7 | 12 |  | Reference Standard | 53 | 9 | 5 |
| Flow and Timing | 51 | 7 | 9 |  |  |  |  |  |

## TABLE OF DESCRIPTION

| **AUTHOR** | **CO**<br>**UN**<br>**TRY**<br>**OF**<br>**ORI**<br>**GIN** | **YE**<br>**AR** | **REFE**<br>**RENC**<br>**E**<br>**STAN**<br>**DAR**<br>**D** | **SENS**<br>**ITIVI**<br>**TY** | **SPEC**<br>**IFICI**<br>**TY** | **True**<br>**Positi**<br>**ve**<br>**(Tp)** | **False**<br>**Positi**<br>**ve**<br>**(FP)** | **Fals**<br>**e**<br>**Neg**<br>**ativ**<br>**e**<br>**(FN**<br>**)** | **True**<br>**Nega**<br>**tive**<br>**(TN)** | **Tot**<br>**al** |
| --- | --- | --- | --- | --- | --- | --- | --- | --- | --- | --- |
| Acharya(6) | Singapore | 2011 | FNA biopsy | 0.98 | 1 | 39 | 0 | 1 | 40 | 80 |
| Acharya(7) | Singapore | 2012 | histopathology | 0.98 | 0.99 | 10 | 0 | 0 | 10 | 20 |
| Acharya(8) | Singapore | 2013 | FNA biopsy | 1 | 1 | 25 | 0 | 0 | 5 | 30 |
| Bartolotta(9) | Italy | 2006 | Histology of resected specimens | 0.77 | 1 | 10 | 0 | 3 | 5 | 18 |
| Cantisani(10) | Italy | 2013 | FNA cytology | 0.79 | 0.91 | 15 | 3 | 4 | 31 | 53 |
| Caresio(11) | Italy | 2018 | histopathology | 1 | 1 | 10 | 0 | 0 | 10 | 20 |
| Chen(12) | China | 2016 | histopathology | 0.86 | 0.88 | 119 | 25 | 17 | 158 | 319 |
| Deng(13) | China | 2014 | histopathology | 0.82 | 0.86 | 46 | 18 | 10 | 101 | 175 |
| Diao(14) | China | 2017 | histopathology | 0.93 | 0.81 | 51 | 6 | 4 | 26 | 87 |
| Ferrari(15) | Italy | 2008 | FNA cytology | 1 | 0.71 | 9 | 2 | 0 | 5 | 16 |
| Friedrich - Rust(16) | Germany | 2009 | FNA cytology | 0.86 | 0.87 | 6 | 6 | 1 | 40 | 53 |
| Giusti M(17) | Italy | 2013 | FNA biopsy | 0.71 | 0.69 | 12 | 5 | 5 | 11 | 33 |
| He(18) | China | 2018 | histopathology | 0.79 | 0.92 | 23 | 5 | 6 | 54 | 88 |
| Jia(19) | China | 2016 | Pathological Diagnosis | 0.8 | 0.83 | 26 | 2 | 2 | 10 | 40 |
| Jiang(20) | China | 2014 | histopathology | 0.97 | 0.95 | 60 | 3 | 2 | 57 | 122 |
| Jiang(21) | China | 2015 | histopathology | 0.9 | 0.92 | 44 | 6 | 5 | 67 | 122 |
| Jin(22) | China | 2017 | histopathology | 0.72 | 0.83 | 26 | 7 | 10 | 34 | 77 |
| Ke | China | 2017 | Pathological Diagnosis | 0.66 | 0.92 | 21 | 3 | 11 | 34 | 69 |
| Li(23) | China | 2013 | FNA biopsy | 0.94 | 0.58 | 199 | 41 | 13 | 57 | 310 |
| Li(24) | China | 2015 | histopathology | 0.88 | 0.8 | 44 | 6 | 6 | 24 | 80 |
| Li(25) | China | 2017 | Pathological Diagnosis | 0.93 | 0.88 | 52 | 4 | 4 | 29 | 89 |
| Liang(26) | China | 2014 | histopathology | 1 | 0.79 | 22 | 12 | 0 | 46 | 80 |
| Liu(27) | China | 2017 | Pathological Diagnosis | 0.6 | 0.89 | 34 | 7 | 23 | 58 | 122 |
| Ma(28) | China | 2014 | histopathology | 0.9 | 0.91 | 85 | 7 | 9 | 71 | 172 |
| Ma X(29) | China | 2017 | Pathological Diagnosis | 0.89 | 0.95 | 70 | 3 | 9 | 53 | 135 |
| Nemec U(30) | Austria | 2012 | Histology of resected specimens | 0.62 | 0.9 | 8 | 3 | 5 | 26 | 42 |
| Schleder (31) | Germany | 2015 | histopathology | 0.92 | 0.81 | 24 | 14 | 2 | 61 | 101 |
| Sui(32) | China | 2016 | histopathology | 0.82 | 0.91 | 54 | 4 | 12 | 39 | 109 |
| Tian(33) | China | 2018 | histopathology | 0.86 | 0.91 | 50 | 9 | 8 | 95 | 162 |
| Wang(34) | China | 2018 | histopathology | 1 | 0.73 | 47 | 24 | 0 | 64 | 135 |
| Wendl(3) | Germany | 2016 | histopathology | 0.8 | 0.63 | 16 | 11 | 4 | 19 | 50 |
| 5) | y |  | gy |  |  |  |  |  |  |  |
| Wu(36) | China | 2015 | FNA or Histology of surgical specimens | 0.75 | 0.85 | 3 | 14 | 1 | 78 | 96 |
| Wu(37) | China | 2016 | FNA biopsy Histology or surgical specimens | 0.95 | 0.67 | 81 | 16 | 4 | 32 | 133 |
| Xu(38) | China | 2019 | histopathology | 0.86 | 0.83 | 221 | 29 | 37 | 145 | 432 |
| Yuan(39) | China | 2015 | histopathology | 0.95 | 0.88 | 35 | 5 | 2 | 36 | 78 |
| Zhang(40) | China | 2016 | histopathology | 0.88 | 0.65 | 81 | 43 | 1 | 32 | 157 |
| Zhang(41) | China | 2017 | Pathological Diagnosis | 0.85 | 0.68 | 46 | 18 | 8 | 39 | 111 |
| Zhang(42) | China | 2018 | histopathology | 0.98 | 0.99 | 41 | 1 | 1 | 77 | 120 |
| Zhang B(43) | China | 2010 | Pathological Diagnosis | 0.88 | 0.92 | 45 | 4 | 6 | 49 | 104 |
| Zhang | China | 2008 | Pathol | 0.6 | 0.91 | 16 | 1 | 9 | 10 | 36 |
| XF(44) | a |  | ogical<br>Diagn<br>osis |  |  |  |  |  |  |  |
| Zhang<br>Y(45) | Chin<br>a | 2017 | FNA<br>biopsy<br>or<br>Histol<br>ogy or<br>surgic<br>al<br>specim<br>ens | 0.77 | 0.94 | 58 | 15 | 17 | 229 | 319 |
| Zhao(46) | Chin<br>a | 2015 | histop<br>atholo<br>gy | 0.81 | 0.82 | 51 | 7 | 12 | 32 | 102 |
| Zhao(47) | Chin<br>a | 2018 | histop<br>atholo<br>gy | 0.82 | 0.71 | 178 | 44 | 39 | 106 | 367 |
| Zhao(48) | Chin<br>a | 2019 | histop<br>atholo<br>gy | 0.89 | 0.88 | 51 | 7 | 6 | 53 | 117 |
| Zhou(49<br>) | Chin<br>a | 2013 | FNA<br>biopsy<br>or<br>Histol<br>ogy or<br>surgic<br>al<br>specim<br>ens | 0.93 | 0.91 | 64 | 10 | 5 | 100 | 179 |
| Zhou(50<br>) | Chin<br>a | 2018 | histop<br>atholo<br>gy | 0.8 | 0.8 | 78 | 14 | 19 | 56 | 167 |

## DISCUSSION

In this systematic review and meta-analysis including 46 studies, we analyzed the use of contrast-enhanced ultrasound as a screening tool rather than a diagnostic tool. The results showed that the concordance and specificity values of CEUS for thyroid nodules were 87% and 84%, with an area under the curve (AUC) of 0.84 and a diagnostic odds ratio (DOR) of 34.97. . Furthermore, the accuracy is 85.4%. The best predictive value was 0.83. Contrast-enhanced ultrasound (CEUS) combines microbubble backscatter with nonlinear acoustics, increasing the ability of physicians to gather information about blood flow in wounds. This is accomplished by injecting a foreign substance into the scars and then watching the blood flow into them. By measuring various imaging parameters to detect angiogenesis in lesions, CEUS outperforms other methods in staging and identification. CEUS can monitor real-time, dynamic and continuous intralesional microcirculation, providing greater insight into the growth characteristics of tumor blood vessels compared to color Doppler ultrasound. It has been shown that CEUS can accurately show the pathological microvascular characteristics of nodules, which is very useful for determining the benign and malignant disease of thyroid nodules. Beneficial nodules appear smooth and round, while malignant nodules appear spiculated or lobulated. In general, benign nodules are smaller than 1, but malignant nodules are larger than 1. In benign nodules, they are often focal and widespread., the symptoms are patchy and granular, but in malignant nodules they are variable and localized. . According to CEUS, statistical differences were found in different degrees of enhancement, enhancement of borders and enhancement patterns when differentiating between positive and negative thyroid nodules . In most cases, benign thyroid nodules show a distribution of blood vessels with a regular appearance, the vessels are arranged in the same way, and often there is a capsule. The blood vessels inside are able to repair continuously, and the edges of the lesions show clearness and continuous repair (51). CEUS has the following disadvantages. In the beginning, it lasts for 5 to 10 minutes in the blood when it is removed by the immune cells, the liver or the spleen, giving the process a little time, already define in advance and complete. Second, ultrasound generates more heat at higher frequencies, so adjusting the ultrasound frequency is very important. Finally, micro-bubbles break at low ultrasound frequencies and at higher mechanical indices (MI), indicating negative sound pressure of the ultrasound system. Although increased MI may improve prognosis, it is associated with impaired microbial bleb degradation, which may lead to microvessel damage and hemolysis (52).

## Conclusion

We can conclude from this systematic review and meta-analysis, which includes 46 articles, that contrast-enhanced ultrasound is more useful as a screening method than as a diagnostic method. The results showed that the concordance and specificity values of CEUS for thyroid nodules were 87% and 84%, with an area under the curve (AUC) of 0.84 and a diagnostic odds ratio (DOR) of 34.97. . Furthermore, the accuracy is 85.4%. The best predictive value was 0.83. Subgroup analysis showed that CEUS had a higher diagnostic accuracy for small nodules (≤ 1 cm) than for larger nodules. This may be because CEUS can provide more information about the vascularity of thyroid nodules, which is particularly useful for evaluating small nodules that may be difficult to detect with conventional ultrasound. The meta-analysis also showed that the diagnostic accuracy of CEUS was influenced by the diagnostic criteria used. Studies that used multiple CEUS features to detect malignant thyroid nodules had higher diagnostic accuracy than studies that used a single CEUS feature. This indicates that it is important to use a holistic approach to interpret CEUS images. Overall, the findings of this meta-analysis suggest that CEUS is a valuable tool for the differential diagnosis of benign and malignant thyroid nodules, especially for small nodules. CEUS can be used to identify thyroid nodules at risk of malignancy, thereby reducing the need for unnecessary fine needle aspiration biopsies.However, it is important to note that CEUS is not a complete test. There is considerable overlap in the CEUS features of benign and malignant thyroid nodules. In addition, the diagnostic accuracy of CEUS is influenced by the experience of the operator and the quality of the ultrasound equipment. CEUS is not currently recommended for the evaluation of all thyroid nodules. However, it may be considered for patients with thyroid nodules suspected of being malignant on conventional ultrasound, for patients with thyroid nodules that are difficult to detect on conventional ultrasound. Therefore, CEUS should not be used to diagnose thyroid cancer. It is important to consider all clinical and imaging findings when making a diagnosis..

## Data Availability

All data produced in the present work are contained in the manuscript

## References

1. Zamora EA, Khare S, Cassaro S. Thyroid Nodule. 2024.

2. Abbarah S, Abbarh S, AlHarthi B. Unusual Thyroid Nodule: A Case of Symptomatic Thyroid Schwannoma. Cureus. 2020 Nov 10;12(11):e11425.

3. Tamhane S, Gharib H. Thyroid nodule update on diagnosis and management. Clin Diabetes Endocrinol. 2016 Dec 3;2(1):17.

4. Acharya UR, Faust O, Sree S V., Molinari F, Garberoglio R, Suri JS. Cost-Effective and Non-Invasive Automated Benign & Malignant Thyroid Lesion Classification in 3D Contrast-Enhanced Ultrasound Using Combination of Wavelets and Textures: A Class of ThyroScan Algorithms. Technol Cancer Res Treat. 2011 Aug 1;10(4):371–80.

5. Lee H, Kim H, Han H, Lee M, Lee S, Yoo H, et al. Microbubbles used for contrast-enhanced ultrasound and theragnosis: a review of principles to applications. Biomed Eng Lett. 2017 May 14;7(2):59–69.

6. Lee H, Kim H, Han H, Lee M, Lee S, Yoo H, et al. Microbubbles used for contrast-enhanced ultrasound and theragnosis: a review of principles to applications. Biomed Eng Lett. 2017 May 14;7(2):59–69.

7. Rajendra Acharya U, Vinitha Sree S, Muthu Rama Krishnan M, Molinari F, Garberoglio R, Suri JS. Non-invasive automated 3D thyroid lesion classification in ultrasound: A class of ThyroScan systems. Ultrasonics. 2012 Apr;52(4):508–20.

8. Acharya UR, Sree SV, Swapna G, Gupta S, Molinari F, Garberoglio R, et al. Effect of complex wavelet transform filter on thyroid tumor classification in three-dimensional ultrasound. Proc Inst Mech Eng H. 2013 Mar 11;227(3):284–92.

9. Bartolotta TV, Midiri M, Galia M, Runza G, Attard M, Savoia G, et al. Qualitative and quantitative evaluation of solitary thyroid nodules with contrast-enhanced ultrasound: initial results. Eur Radiol. 2006 Oct 3;16(10):2234–41.

10. Cantisani V, Bertolotto M, Weskott HP, Romanini L, Grazhdani H, Passamonti M, et al. Growing indications for CEUS: The kidney, testis, lymph nodes, thyroid, prostate, and small bowel. Eur J Radiol. 2015 Sep;84(9):1675–84.

11. Cantisani V, Bertolotto M, Weskott HP, Romanini L, Grazhdani H, Passamonti M, et al. Growing indications for CEUS: The kidney, testis, lymph nodes, thyroid, prostate, and small bowel. Eur J Radiol. 2015 Sep;84(9):1675–84.

12. Chen M, Zhang KQ, Xu YF, Zhang SM, Cao Y, Sun WQ. Shear wave elastography and contrast-enhanced ultrasonography in the diagnosis of thyroid malignant nodules. Mol Clin Oncol. 2016 Dec;5(6):724–30.

13. Deng J, Zhou P, Tian S Ming, Zhang L, Li J le, Qian Y. Comparison of Diagnostic Efficacy of Contrast-Enhanced Ultrasound, Acoustic Radiation Force Impulse Imaging, and Their Combined Use in Differentiating Focal Solid Thyroid Nodules. PLoS One. 2014 Mar 3;9(3):e90674.

14. Diao X, Zhan J, Chen L, Chen Y, Liu Y. Quantification of solid hypo-echoic thyroid nodule enhancement with contrast-enhanced ultrasound. Transl Cancer Res. 2017 Dec;6(6):1078–87.

15. Ferrari FS, Megliola A, Scorzelli A, Guarino E, Pacini F. Ultrasound examination using contrast agent and elastosonography in the evaluation of single thyroid nodules: Preliminary results. J Ultrasound. 2008 Jun;11(2):47–54.

16. Friedrich-Rust M, Sperber A, Holzer K, Diener J, Grünwald F, Badenhoop K, et al. Real-time Elastography and Contrast-Enhanced Ultrasound for the Assessment of Thyroid Nodules. Experimental and Clinical Endocrinology & Diabetes. 2009 Oct 23;118(09):602–9.

17. Giusti M, Orlandi D, Melle G, Massa B, Silvestri E, Minuto F, et al. Is there a real diagnostic impact of elastosonography and contrast-enhanced ultrasonography in the management of thyroid nodules? J Zhejiang Univ Sci B. 2013 Mar 10;14(3):195–206.

18. He Y, Wang XY, Hu Q, Chen XX, Ling B, Wei HM. Value of Contrast-Enhanced Ultrasound and Acoustic Radiation Force Impulse Imaging for the Differential Diagnosis of Benign and Malignant Thyroid Nodules. Front Pharmacol. 2018 Nov 27;9.

19. Zhan J, Diao XH, Chen L, Jin JM, Chen Y. Role of Contrast-Enhanced Ultrasound in Diagnosis of Thyroid Nodules in Acoustic Radiation Force Impulse “Gray Zone.” Ultrasound Med Biol. 2017 Jun;43(6):1179–86.

20. Jiang J, Shang X, Zhang H, Ma W, Xu Y, Zhou Q, et al. Correlation Between Maximum Intensity and Microvessel Density for Differentiation of Malignant From Benign Thyroid Nodules on Contrast-Enhanced Sonography. Journal of Ultrasound in Medicine. 2014 Jul;33(7):1257–63.

21. Jiang J, Shang X, Wang H, Xu Y, Gao Y, Zhou Q. Diagnostic value of contrast-enhanced ultrasound in thyroid nodules with calcification. Kaohsiung J Med Sci. 2015 Mar 17;31(3):138– 44.

22. Jin L, Xu C, Xie X, Li F, Lv X, Du L. An Algorithm of Image Heterogeneity with Contrast-Enhanced Ultrasound in Differential Diagnosis of Solid Thyroid Nodules. Ultrasound Med Biol. 2017 Jan;43(1):104–10.

23. LI F, LUO H. Comparative study of thyroid puncture biopsy guided by contrast-enhanced ultrasonography and conventional ultrasound. Exp Ther Med. 2013 May;5(5):1381–4.

24. LI F, ZHANG J, WANG Y, LIU L. Clinical value of elasticity imaging and contrast-enhanced ultrasound in the diagnosis of papillary thyroid microcarcinoma. Oncol Lett. 2015 Sep;10(3):1371–7.

25. LI F, ZHANG J, WANG Y, LIU L. Clinical value of elasticity imaging and contrast-enhanced ultrasound in the diagnosis of papillary thyroid microcarcinoma. Oncol Lett. 2015 Sep;10(3):1371–7.

26. Liang XN, Guo RJ, Li S, Zheng ZM, Liang HD. Binary logistic regression analysis of solid thyroid nodules imaged by high-frequency ultrasonography, acoustic radiation force impulse, and contrast-enhanced ultrasonography. Eur Rev Med Pharmacol Sci. 2014;18(23):3601–10.

27. Liu MJ, Men YM, Zhang YL, Zhang YX, Liu H. Improvement of diagnostic efficiency in distinguishing the benign and malignant thyroid nodules via conventional ultrasound combined with ultrasound contrast and elastography. Oncol Lett. 2017 Jul;14(1):867–71.

28. Ma J jiao, Ding H, Xu B hua, Xu C, Song L jun, Huang B jian, et al. Diagnostic Performances of Various Gray-Scale, Color Doppler, and Contrast-Enhanced Ultrasonography Findings in Predicting Malignant Thyroid Nodules. Thyroid. 2014 Feb;24(2):355–63.

29. Ma X, Zhang B, Ling W, Liu R, Jia H, Zhu F, et al. Contrast-enhanced sonography for the identification of benign and malignant thyroid nodules: Systematic review and meta-analysis. Journal of Clinical Ultrasound. 2016 May 24;44(4):199–209.

30. Nemec U, Nemec SF, Novotny C, Weber M, Czerny C, Krestan CR. Quantitative evaluation of contrast-enhanced ultrasound after intravenous administration of a microbubble contrast agent for differentiation of benign and malignant thyroid nodules: assessment of diagnostic accuracy. Eur Radiol. 2012 Jun 10;22(6):1357–65.

31. Schleder S, Janke M, Agha A, Schacherer D, Hornung M, Schlitt HJ, et al. Preoperative differentiation of thyroid adenomas and thyroid carcinomas using high-resolution contrast-enhanced ultrasound (CEUS). Clin Hemorheol Microcirc. 2015 Oct 28;61(1):13–22.

32. Sui X, Liu HJ, Jia HL, Fang QM. Contrast-enhanced ultrasound and real-time elastography in the differential diagnosis of malignant and benign thyroid nodules. Exp Ther Med. 2016 Aug;12(2):783–91.

33. Tian Q, Zhu H, Li H. Significance of contrast-enhanced ultrasonography in the differential diagnosis of thyroid nodules. Medicine. 2018 Oct;97(40):e12688.

34. Wang Y, Nie F, Liu T, Yang D, Li Q, Li J, et al. Revised Value of Contrast-Enhanced Ultrasound for Solid Hypo-Echoic Thyroid Nodules Graded with the Thyroid Imaging Reporting and Data System. Ultrasound Med Biol. 2018 May;44(5):930–40.

35. Wendl CM, Janke M, Jung W, Stroszczysnski C, Jung EM. Contrast-enhanced ultrasound with perfusion analysis for the identification of malignant and benign tumors of the thyroid gland. Clin Hemorheol Microcirc. 2016 Aug 27;63(2):113–21.

36. Wu Q, Wang Y, Li Y, Hu B, He ZY. Diagnostic value of contrast-enhanced ultrasound in solid thyroid nodules with and without enhancement. Endocrine. 2016 Aug 5;53(2):480–8.

37. Wu Q, Li Y, Wang Y. Diagnostic value of “absent” pattern in contrast-enhanced ultrasound for the differentiation of thyroid nodules. Clin Hemorheol Microcirc. 2016 Oct 5;63(4):325–34.

38. Xu Y, Qi X, Zhao X, Ren W, Ding W. Clinical diagnostic value of contrast-enhanced ultrasound and TI-RADS classification for benign and malignant thyroid tumors. Medicine. 2019 Jan;98(4):e14051.

39. Yuan Z, Quan J, Yunxiao Z, Jian C, Zhu H. Contrast-enhanced ultrasound in the diagnosis of solitary thyroid nodules. J Cancer Res Ther. 2015;11(1):41.

40. Zhang Y, Luo Y Kun, Zhang M bo, Li J, Li J, Tang J. Diagnostic Accuracy of Contrast-Enhanced Ultrasound Enhancement Patterns for Thyroid Nodules. Medical Science Monitor. 2016 Dec 5;22:4755–64.

41. Zhang Y, Luo Y Kun, Zhang M bo, Li J, Li J, Tang J. Diagnostic Accuracy of Contrast-Enhanced Ultrasound Enhancement Patterns for Thyroid Nodules. Medical Science Monitor. 2016 Dec 5;22:4755–64.

42. Zhang Y, Luo Y Kun, Zhang M bo, Li J, Li J, Tang J. Diagnostic Accuracy of Contrast-Enhanced Ultrasound Enhancement Patterns for Thyroid Nodules. Medical Science Monitor. 2016 Dec 5;22:4755–64.

43. Zhang Y, Luo Y Kun, Zhang M bo, Li J, Li J, Tang J. Diagnostic Accuracy of Contrast-Enhanced Ultrasound Enhancement Patterns for Thyroid Nodules. Medical Science Monitor. 2016 Dec 5;22:4755–64.

44. Zhang Y, Luo Y Kun, Zhang M bo, Li J, Li J, Tang J. Diagnostic Accuracy of Contrast-Enhanced Ultrasound Enhancement Patterns for Thyroid Nodules. Medical Science Monitor. 2016 Dec 5;22:4755–64.

45. Zhang Y, Zhou P, Tian SM, Zhao YF, Li JL, Li L. Usefulness of combined use of contrast-enhanced ultrasound and TI-RADS classification for the differentiation of benign from malignant lesions of thyroid nodules. Eur Radiol. 2017 Apr 15;27(4):1527–36.

46. Zhang Y, Zhou P, Tian SM, Zhao YF, Li JL, Li L. Usefulness of combined use of contrast-enhanced ultrasound and TI-RADS classification for the differentiation of benign from malignant lesions of thyroid nodules. Eur Radiol. 2017 Apr 15;27(4):1527–36.

47. Zhang Y, Zhou P, Tian SM, Zhao YF, Li JL, Li L. Usefulness of combined use of contrast-enhanced ultrasound and TI-RADS classification for the differentiation of benign from malignant lesions of thyroid nodules. Eur Radiol. 2017 Apr 15;27(4):1527–36.

48. Zhang Y, Zhou P, Tian SM, Zhao YF, Li JL, Li L. Usefulness of combined use of contrast-enhanced ultrasound and TI-RADS classification for the differentiation of benign from malignant lesions of thyroid nodules. Eur Radiol. 2017 Apr 15;27(4):1527–36.

49. Zhou Q, Xu YB, Jiang J, Ma WQ, Wang H, Li M, et al. [Differential diagnostic value of contrast-enhanced ultrasound in calcified thyroid nodules]. Zhonghua Er Bi Yan Hou Tou Jing Wai Ke Za Zhi. 2013 Sep;48(9):726–9.

50. Zhou X, Zhou P, Hu Z, Tian SM, Zhao Y, Liu W, et al. Diagnostic Efficiency of Quantitative Contrast-Enhanced Ultrasound Indicators for Discriminating Benign From Malignant Solid Thyroid Nodules. Journal of Ultrasound in Medicine. 2018 Feb 7;37(2):425–37.

51. Zhou X, Zhou P, Hu Z, Tian SM, Zhao Y, Liu W, et al. Diagnostic Efficiency of Quantitative Contrast-Enhanced Ultrasound Indicators for Discriminating Benign From Malignant Solid Thyroid Nodules. Journal of Ultrasound in Medicine. 2018 Feb 7;37(2):425–37.

52. Zhan J, Ding H. Application of contrast-enhanced ultrasound for evaluation of thyroid nodules. Ultrasonography. 2018 Oct 1;37(4):288–97.

